# Pediatric Spigelian Hernia and Spigelian-Cryptorchidism Syndrome: an Integrating Review

**DOI:** 10.1101/2025.01.24.25321083

**Authors:** Javier Arredondo Montero, María Rico-Jiménez

## Abstract

**Introduction:** Spigelian hernia (SH) is an aponeurotic defect, either acquired or congenital, in Spiegel’s semilunar line. SH is exceptional in pediatric patients.

**Methods:** A comprehensive non-systematic review of the previous literature was conducted. Eligible studies were identified by searching the primary existing medical bibliography databases. Median and interquartile range or mean and standard deviation were used to describe quantitative variables and proportions for categorical variables. The Kruskal-Wallis, Mann-Whitney U, and Fisher’s exact tests were used to compare group variables. The Spearman, Pearson and Cramér’s V correlation analyses were used to assess the degree of correlation between the study variables. A p-value <0.05 (two tails) was considered statistically significant.

**Results:** Eighty-two publications reporting 123 patients were included. Of these, 105 were male (85.4%). The age range was from 0 to 21 years. Forty-seven patients (38.2%) had a left-sided SH, and 13 (10.6%) had a bilateral SH. Forty-five cases (36.6%) were classified as traumatic, the majority attributable to bicycle-related injuries. Forty-one patients (33.3%) presented with a SH associated with undescended testis (UDT). A peak incidence around 7-9 years was identified for traumatic SH, and a concentration of cases before one year of age for SH associated with UDT. Fifteen patients (12.2%) were reported to have hernia incarceration/strangulation (I/S). These patients were significantly younger than those without I/S (p=0.02), but no gender differences were seen (p=0.63). In 95 patients (77.2%), surgical correction of the defect was reported. Fourteen were approached laparoscopically, with a 35.7% conversion rate. Eight (6.5%) were managed conservatively. Overall, the reported evolution has been favorable.

**Conclusions:** HS is an uncommon condition in pediatric populations, predominantly affecting males. It can present congenitally, with a significant association with ipsilateral UDT, or it can be acquired, typically related to bicycle trauma involving the SL. The risk of incarceration is relatively high, particularly during early childhood. Most reported cases have been treated surgically, with favorable outcomes. Evidence regarding conservative management is limited.

## Introduction

Abdominal wall hernias are one of the most common pediatric surgical pathologies worldwide. However, the incidence of the different types of hernia are highly variable. While inguinal and umbilical hernias are frequent, other types, such as femoral or Spigelian hernias (SH), are highly infrequent [1].

### Anatomy and Embryology of Semilunaris Line (Spigelian Line)

During embryonic development, the primitive mesoderm formed during gastrulation differentiates into the splanchnic mesoderm (which develops into the peritoneum) and the somatic mesoderm (which contributes to the formation of the abdominal wall). Between the fourth and tenth weeks of gestation, myocytes migrate bilaterally from the paravertebral regions to form the muscular and aponeurotic layers of the anterior abdominal wall, completing their fusion and establishing the linea alba by approximately week ten. In the following weeks, these layers gain strength, and the superficial fascia undergoes complete differentiation [6–8]. This process is often associated with congenital points of weakness, such as the linea alba (where epigastric hernias occur) or the umbilicus (where umbilical hernias develop).

The *linea semilunaris* or Spigelian line (SL) was first described by Adriaan Van Den Spiegel (1578-1625) as the region of the anterior abdominal wall formed by the transition from muscle to aponeurosis of the *transversus abdominis* muscle of the abdomen. It is a structure located on the lateral margin of the *anterior rectus abdominis* muscle. It extends from the inferior edge of the costal cartilage (approximately at the ninth rib) to the pubic tubercle, following a curved and descending path along the anterior abdominal wall. It is considered a point of fascial weakness [1–3]. Although SL partially overlaps within the lateral margin of the *anterior rectus abdominis* muscle, recent studies suggest that the anatomic boundaries of these two structures are different [8].

SL is an infrequent but well-described site where hernias can occur. Hernias that occur along the SL are referred to as SH [4]. These hernias are usually located deep to the external oblique muscle. They are referred to as interparietal hernias (abdominal hernias in which the hernial sac is situated between the layers of the abdominal wall, specifically between the fascias or muscles, rather than completely protruding through them). About pediatric SH, two variants have been described: congenital SH and acquired SH. Figure 1 presents an schematic diagram of the two subtypes of Spigelian hernia (SH) described in the pediatric population (congenital and acquired) and their distinguishing characteristics. The aim of this integrative review is to analyze the existing literature on this condition in the pediatric population to better characterize it in terms of epidemiology, diagnosis, treatment, and prognosis.

**Figure 1.**
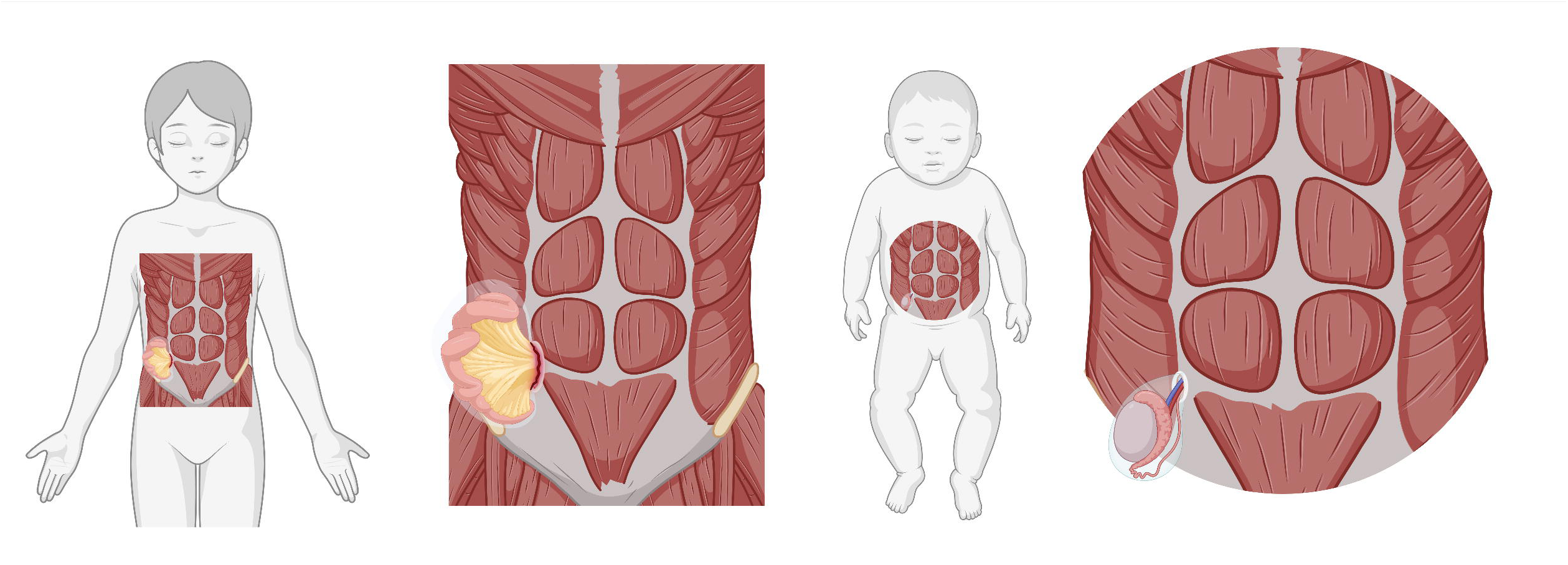
Graphic representation of the two main subtypes of Spiegel’s hernia in the pediatric age group. <u>Left</u>: Acquired (traumatic) Spiegel’s hernia. <u>Right</u>: Spiegel’s hernia in the context of Spigelian-Cryptorchidism Syndrome. Created in BioRender. Arredondo montero, J. (2025) https://BioRender.com/a53g564

## Methods

An integrative non-systematic review of the previous literature was conducted. Eligible studies were identified by searching the primary existing medical bibliography databases (PubMed, Web of Science, Scopus, and OVID). Search terms and keywords were: “(Spiegel OR Spigelian OR semilunar OR semilunaris) AND (hernia) AND (cryptorchidism OR undescended testis OR) AND (pediatric OR children OR infant). Supplementary File 1 shows inclusion and exclusion criteria. JAM and MRJ selected articles using the COVIDENCE ® tool. The search results were imported into the platform, and both authors screened the articles separately. Disagreements were resolved by consensus.

Regarding descriptive statistics, median and interquartile range or the mean and standard deviation were used for quantitative variables and proportions for categorical variables. The Kolmogorov-Smirnov test was applied to assess the normality of quantitative variables. Mann-Whitney U, Kruskal-Wallis, and Fisher’s exact tests were used to compare sociodemographic and clinical variables between groups. Pearson, Spearman, and Cramér’s V correlation analyses were performed to assess the degree of correlation between the study variables. A p-value of <0.05 (two-tailed) was considered statistically significant. All statistical analyses were conducted using STATA 18.0 (StataCorp LLC). Graphical plots were generated using Python (version 3.9). The visualizations were created with the Seaborn library (version 0.11.2) for histograms and kernel density estimates and Matplotlib (version 3.4.3) for plotting. Kernel density plots were generated using default bandwidth parameters, which were adjusted to ensure smoothness in the density curves. Supplementary file 2 includes a database with the main variables analyzed.

## Results

### Main Characteristics of the Included Studies. Sociodemographic Features of the Patients

The literature review, covering the PubMed/Medline, Web of Science, Scopus, and OVID databases, identified 82 indexed publications by 79 authors [9–90]. Table 1 summarizes all the published cases of congenital Spigelian hernia (SH) described in the pediatric population to date.

The distribution of publications by country was as follows: USA (n=23), India (n=10), Italy (n=6), Turkey (n=5), Spain (n=4), United Kingdom (n=4), Australia (n=4), Greece (n=3), Israel (n=3), Japan (n=3), New Zealand (n=2), and Saudi Arabia (n=2). Algeria, Ireland, Norway, Germany, Puerto Rico, Canada, Ghana, and Tunisia each contributed one publication. The geographic origin of five publications could not be determined. A total of 123 patients were reported. Of these, 105 were male (85.4%) and 18 were female (14.6%). The age range was from 0 days to 21 years (mean = 5.21, standard deviation = 5.34). Forty-six patients were one year or younger.

### Characteristics of the Spigelian Hernia

Forty-seven patients (38.2%) had a left-sided SH, 56 (45.5%) had a right-sided SH, and 13 (10.6%) had a bilateral SH. In seven cases (5.7%) laterality was not explicitly reported. Concerning SH measurements, 55 cases reported numerical values, of which 15 provided two independent values in centimeters (major and minor axes), and 40 provided a single value in centimeters (corresponding to the major axis). A significant heterogeneity was identified in the methodology used to determine the measurements. Some authors utilized radiological or intraoperative measurements of the fascial defect (ring), while others, such as Fraser et al., measured the diameter of the cutaneous bulge [40]. Some authors, such as Vaos et al., Inan et al., and Upasani et al., provided both characterizations, noting a substantially larger size of the cutaneous protrusions compared to the actual size of the fascial defect [44,56,65].On the other hand, some authors, such as Bilici et al. and Okumuş et al. [87], provided a range of measurements for the defect [60,87]. For the statistical analysis of these patients, the central value of the range was used as the imputed value.

For cases reported as a single measurement, the mean (standard deviation) was 3.21 (2.23) cm. For cases reported with two measurements, the values were converted to surface area (cm²), resulting in a mean (standard deviation) of 19.41 (24.37) cm². No statistically significant differences were found in the size of fascial defects between patients with undescended testis (UDT) and those without UDT (p=0.96), nor between patients with traumatic etiology and those without traumatic etiology (p=0.45). No differences in fascial size were observed based on age (Spearman, p=0.93) or sex (p=0.68). Similarly, no statistically significant differences were found concerning laterality (Kruskal Wallis, p=0.65).

### Spigelian Hernia Etiology and associations

Regarding etiology, 45 out of 123 cases (36.6%) were classified as acquired or traumatic. Specifically, 2 cases (1.6%) were caused by unspecified local trauma, 32 cases (26%) by bicycle (n=31) or motorcycle (n=1) handlebar trauma, 2 cases (1.6%) by bicycle falls, 1 case (0.8%) by an ATV accident, 2 cases (1.6%) by BMX accidents, 1 case (0.8%) by a nail puncture, 1 case (0.8%) by cow goring, 2 cases (1.6%) by road or automobile accidents, and 3 cases (2.4%) related to surgical procedures (1 case of Bochdalek hernia associated with intense parietal traction during surgical correction, 1 case of bilateral inguinal herniorrhaphy with hernia detected in the immediate postoperative period, and 1 case of a surgically treated lower mediastinal neuroblastoma). The remaining cases were either congenital or had no attributable etiology described. No statistically significant differences were found between patients with traumatic and non-traumatic SH based on gender (p=0.57).

Forty-one patients (33.3%) presented with a SH associated with UDT. Concerning laterality, 16 (39%) UDT were left-sided, 15 (36.6%) right-sided, and 10 (24.4%) bilateral. Fisher’s exact test confirmed a statistically significant association between the side of SH and the side of UDT (p<0.0001). When a Cramér’s V analysis was performed between the side of SH and the side of UDT, a very strong association was found (0.83).

Regarding age and the etiology of SH, the mean age of patients with traumatic SH was 9.2 (4) years, while for patients with non-traumatic SH, it was 2.9 (4.6) years (p<0.0001). Similarly, patients with SH and associated UDT had a mean age of 0.7 (1.1) years, compared to a mean age of 7.5 (5.2) years for patients without UDT (p<0.0001).

Histograms and kernel density plots (Figure 2) were used to visually represent both etiologies, highlighting a peak incidence of traumatic SH around 7–9 years and a concentration of SH cases associated with UDT occurring predominantly before one year of age. In this case, the presence of UDT was used as a reference for congenital SH in both epidemiological and analytical terms, although a perfect equivalence cańt be assumed.

**Figure 2.**
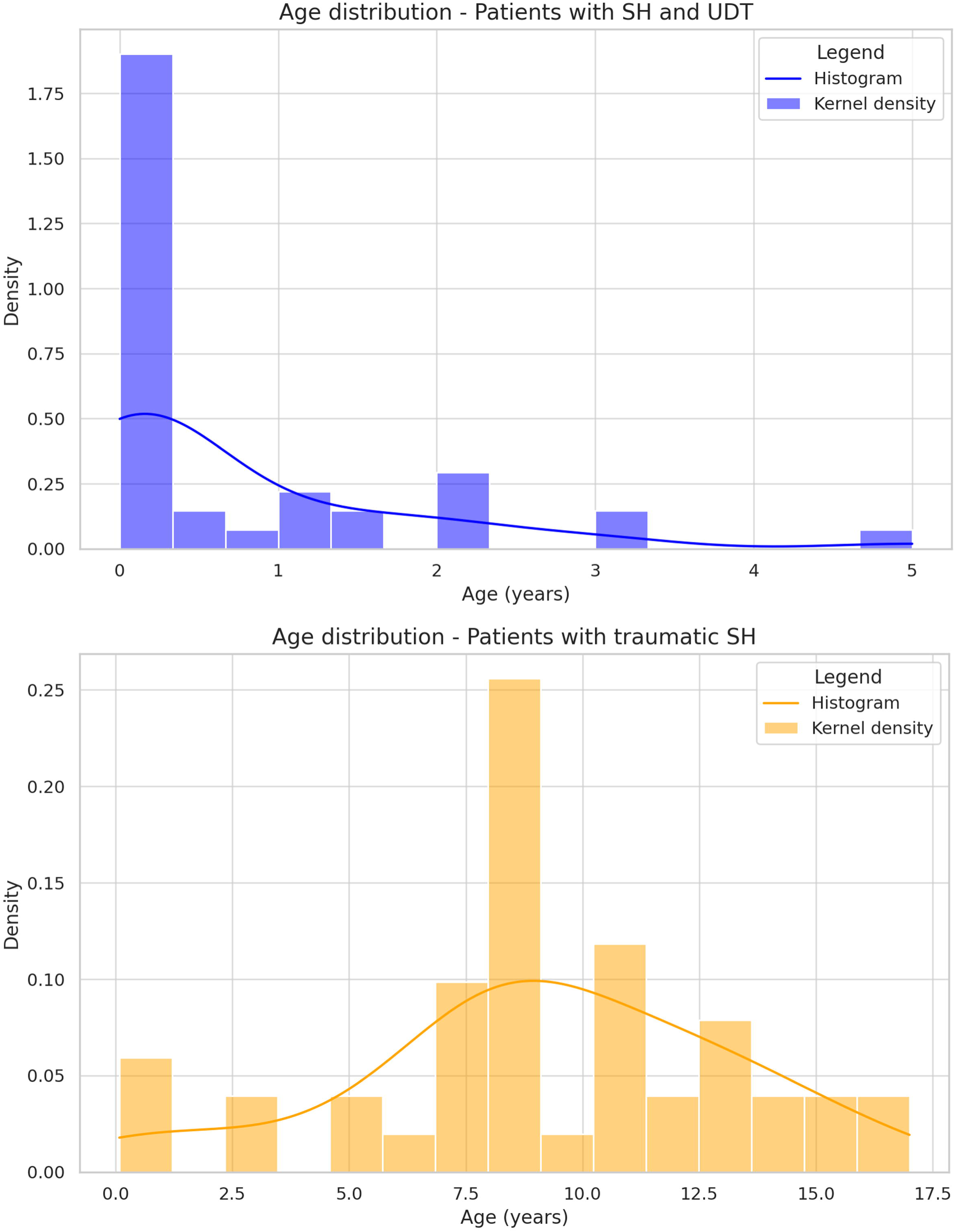
Histogram and kernel density plots showing the age distribution of SH associated with UDT (top) and trauma-related SH (bottom).

Lastly, Table 1 details the associated malformations of patients diagnosed with HS.

### Complications Associated with Spigelian Hernia

Fifteen out of 123 patients (12.2%) were reported to have some hernia incarceration or strangulation. These 15 patients were significantly younger than the group of patients without hernia incarceration: 2.7 (4.3) years vs. 5.6 (5.4) years (p=0.02), but no gender differences were seen (p=0.63). Additionally, no differences were found in the hernia incarceration rate between the UDT group and the non-UDT group (p=0.37) nor between the traumatic and non-traumatic SH groups (p=0.25). Seven visceral injuries (5.6%) were described, all of them in patients with traumatic SH and attributable to the injury mechanism that caused the SH (p=0.001).

### Spigelian Hernia Diagnosis

Regarding the most commonly used imaging modalities for the diagnosis of SH, plain abdominal X-rays were reported in 10 patients (8.1%), with most cases corresponding to traumatic or incarcerated/strangulated SH. In 32 patients (26%), ultrasound scan (US) was reported as the imaging modality used, and in 20 patients (16.3%), computed tomography (CT) was reported, with the vast majority being trauma cases. Several authors reported using two or more imaging modalities.

### Spigelian Hernia Treatment

In 95 patients (77.2%), surgical correction of the defect was reported, and in at least 15 of them (15.8%), it was performed urgently. The treatment provided was not detailed in 9 studies (9.6%). Of the surgically treated patients, 14 (14.7%) were approached laparoscopically. Of these, 8 (57.1%) were repaired entirely laparoscopically, 1 (7.1%) was laparoscopically explored but not repaired, and 5 (35.7%) required conversion to open surgery.

In most cases, primary repair of the defect was performed. Few authors reported the use of mesh for the repair of pediatric SH. Singal et al. [55] reported using a VYPRO™ mesh and Fascetti-Leon et al. reported using a Vicryl™ mesh [51].

Eight patients (6.5%) were managed conservatively. Of these, 3 (2.4%) had a complete spontaneous resolution, 3 (2.4%) had a partial resolution (these 3 cases were traumatic SH with a limited follow-up period), 1 (0.8%) patient died, and 1 (0.8%) had no reported follow-up.

Table 1 provides specific details for each case regarding the surgical approach, findings, and the available follow-up period.

### Outcomes. Complications Associated with Spigelian Hernia corrective surgery

Ninety-five patients (77.2%) were reported as having a favorable outcome, with a highly variable follow-up period. Sixteen (13%) had no data on their follow-up and outcome. One of the patients (0.8%) died due to SH strangulation. Concerning post-surgical complications, one patient (0.8%) experienced persistent postoperative pain, 1 (0.8%) had abdominal wall laxity, and 3 patients with SH associated with UDT (2.4%) developed a scrotal abscess. Two of them developed testicular atrophy secondary to the process.

## Discussion

The reported cases demonstrate a broad geographical distribution and no consistent pattern was identified. We found a clear predominance of this pathology in males (85.4%) and a slight predominance of right-sided HS over left-sided HS (45.5% vs. 38.2%), with a relatively high proportion of bilateral HS cases (10.6%). A significant variability has been identified in the way fascial defects are characterized, making it challenging to extrapolate information on this aspect.

Trauma-related HS accounts for 37.2% of reported cases in the literature. This type predominantly occurs in the age range of 7 to 9 years, with no sex predominance. Trauma-related HS has been associated with various injury mechanisms, with a clear predominance of activities involving bicycles, motorcycles, and BMX bikes. The most common mechanisms are sudden deceleration and blunt impact of the handlebar on the SL. In these cases, visceral injuries of various kinds have been documented. Therefore, in the presence of trauma with associated SH (or handlebar trauma), one must assume a high-energy injury mechanism and conduct a thorough and exhaustive screening for underlying injuries that could compromise life and whose diagnostic delay may be associated with morbidity and mortality. It is also relevant to mention that several cases have been reported as ‘handlebar hernia’ in a nonspecific manner. In our literature review, we were able to categorize some of these as SH (based on anatomical description or CT images), but others corresponded to anatomical areas of the abdominal wall different from SL. For this reason, we consider it essential that future authors report the anatomy of the lesions accurately and in detail, as there may be diagnostic, therapeutic, and prognostic variations between SH and other parietal lesions.

On the other hand, SH associated with UDT represents 32.2% of pediatric SH reported cases. In most instances, this is assumed to be a congenital hernia, with a strong correlation between the side of the UDT and the hernia. It is striking the high number of patients with associated anterior wall defects, ranging from non-formed inguinal canals to hernias. For example, Silberstein et al. [32] reported an absence of inguinal canal formation with associated muscle atrophy. Ostlie et al. [35], Raveenthiram [43], Bilici et al. [60] and Parihar et al. [63] reported similar findings, and Al-Salem et al. [37], Ostlie et al. [35], Bilici et al. [60] and Parihar et al. [63] reported an absence of *Gubernaculum testis* formation [35]. More recent authors, such as Gonuguntla et al. [85] reported similar findings. In general, when muscular alterations have been reported, they have been described in relation to the internal oblique and transversus abdominis muscles and have been described as hypoplastic, thinned out, or even absent (Komura et al., Singal et al., Inan et al.) [29,55,56]. Komura et al. reported in 1994 [29] the histology of the muscles adjacent to the affected area in HS, demonstrating atrophy with fat infiltration and interstitial fibrosis. In this context, it may be necessary to differentiate between congenital and acquired HS in terms of the likelihood of spontaneous resolution with conservative management (which might be more likely in cases of traumatic HS). A more comprehensive characterization of the defect in future cases could influence the choice of the best therapeutic approach, reserving conservative management for minor asymptomatic non-congenital fascial defects. Nevertheless, the evidence in this regard is limited. Lastly, it is relevant to consider that some authors have proposed that UDT and SH may constitute a pathological spectrum encompassed under the term “Spigelian-Cryptorchidism Syndrome”. It has also been suggested that this could be a variant of Prune-Belly syndrome, although we have not found a consistent pattern of visceral, cardiac, genitourinary, or musculoskeletal malformations in the reported patients.

Regarding complications, a relatively high percentage of patients (12.2%) experienced hernia incarceration or strangulation, with this complication predominantly occurring in younger patients. This percentage, significantly higher than other hernia-related conditions such as umbilical or epigastric hernias, should be considered when recommending prompt correction of the defect, especially during early childhood.

Diagnosing this condition has traditionally been considered challenging due to the limited clinical expression of the defect (secondary to its intraparietal location). Given that pediatric patients have a thinner anterior abdominal wall due to less muscular development and a reduced adipose layer, these hernias are potentially easier to identify clinically. However, familiarity with the anatomy of the abdominal wall and a high index of suspicion are essential for an accurate diagnosis. Several cases of misdiagnosis and diagnostic confusion with other pediatric abdominal wall defects, such as inguinal hernia, have been reported [45].

Concerning radiology tests, this review highlights the use of different imaging modalities for this condition, with a predominance of plain abdominal X-rays and CT scans in urgent or trauma scenarios and an increasing use of ultrasound in elective settings, particularly in recent years. It is important to emphasize that SH is an uncommon diagnosis in the pediatric population, and radiological studies should be performed in a targeted manner by expert radiologists to avoid diagnostic errors.

The most widely accepted and utilized treatment approach in the existing literature is open surgery. Laparoscopy has been employed in selective cases, although it is associated with a relatively high conversion rate. Conservative management is scarcely documented for this condition but has proven effective in some patients. From a perspective of biological plausibility and surgical safety, we believe that conservative management should be limited to small fascial defects in healthy patients with a traumatic etiology, who are asymptomatic and show no herniated contents on imaging studies. Likewise, we consider that these patients require close clinical and ultrasound follow-up.

Concerning the use of mesh, although the evidence is limited, given the favorable healing profile of the pediatric population and the excellent outcomes reported in cases with primary repair, the routine use of mesh for the surgical correction of this condition is not justified.

Regarding the surgical prognosis of UDT specifically in this context, the majority of authors have reported the successful performance of primary orchidopexy in a single stage. We attribute this to the fact that the hernial sac allows for progressive elongation of the spermatic vessels and the vas deferens, in contrast to purely intra-abdominal testes. Concerning medium and long term results, most published cases report a favorable outcome, but with a variable follow-up period. However, in many cases, follow-up is limited or not reported at all. Once again, it is essential to address this aspect to better understand this condition in prognostic terms.

This review has significant strengths, such as the comprehensive analysis of the existing literature and its statistical approach, with findings relevant to understanding the disease. However, it also has notable limitations, including the low number of reported cases of the condition, the lack of data or incomplete reports for some patients, the use of inferential statistics, and the inability to perform meta-analytical models.

In conclusion, HS is an uncommon condition in pediatric populations, predominantly affecting males. It can present congenitally, with a significant association with ipsilateral UDT, or it can be acquired, typically related to bicycle trauma involving the SL. The risk of incarceration is relatively high, particularly during early childhood. Most reported cases have been treated surgically, with favorable outcomes. Evidence regarding conservative management is limited.

## Contributor declaration page (CRediT statement)

Dr. Javier Arredondo Montero conceptualized and designed the study, collected and analyzed the data, drafted the initial manuscript, and revised the manuscript.

Dr. María Rico-Jiménez collected the data and revised the manuscript.

## Original work

All manuscript authors declare this is an original contribution that has not been previously published.

## Data availability

The dataset used to carry out this study is attached as Supplementary File 2.

## Informed consent

N/A.

## Ethical Approval

This study is a review of the literature in which neither human nor animal patients participated.

Therefore, no ethics committee assessment was sought.

## References

[1] Skandalakis PN, Zoras O, Skandalakis JE, Mirilas P. Spigelian hernia: surgical anatomy, embryology, and repair technique. Am Surg. 2006 Jan;72(1):42–8. PMID: 16494181.

[2] Huttinger R, Sugumar K, Baltazar-Ford KS. Spigelian Hernia. 2023 Jun 26. In: StatPearls [Internet]. Treasure Island (FL): StatPearls Publishing; 2024 Jan–. PMID: 30855874.

[3] Hanzalova I, Schäfer M, Demartines N, Clerc D. Spigelian hernia: current approaches to surgical treatment-a review. Hernia. 2022 Dec;26(6):1427–1433. doi: 10.1007/s10029-021-02511-8. Epub 2021 Oct 19. PMID: 34665343; PMCID: PMC9684297.

[4] Jones BC, Hutson JM. The syndrome of Spigelian hernia and cryptorchidism: a review of paediatric literature. J Pediatr Surg. 2015 Feb;50(2):325–30. doi: 10.1016/j.jpedsurg.2014.10.059. Epub 2014 Nov 5. PMID: 25638630.

[5] Larsen, W. J. (2014). Human Embryology. Elsevier.

[6] Sadler, T. W. (2020). Langman’s Medical Embryology. Wolters Kluwer.

[7] Moore, K. L., Persaud, T. V. N., & Torchia, M. G. (2020). Before We Are Born: Essentials of Embryology and Birth Defects. Elsevier.

[8] Vierstraete M, Pereira Rodriguez JA, Renard Y, Muysoms F. EIT Ambivium, Linea Semilunaris, and Fulcrum Abdominalis. J Abdom Wall Surg. 2023 Dec 22;2:12217. doi: 10.3389/jaws.2023.12217. PMID: 38312427; PMCID: PMC10831682.

[9] Scopinaro AJ: Hernia in Spigel’s semilunar line in a newborn. Semana Med 1:284.1935

[10] Hurwitt ES, Borow M. Bilateral Spigelian hernias in childhood. Surgery. 1955 Jun;37(6):963–8. PMID: 14373320.

[11] Landry RM. Traumatic hernia. Am J Surg. 1956 Feb;91(2):301–2. doi: 10.1016/0002-9610(56)90427-5. PMID: 13283220.

[12] Isaacson NH (1956) Spigelian Hernia. Report of four cases. Med Ann Dist Columbia 58: 23–26

[13] Wilson TH Jr. Traumatic hernia of the abdominal wall. Am J Surg. 1959 Mar;97(3):340–1. doi: 10.1016/0002-9610(59)90314-9. PMID: 13627364.

[14] Roberts GR. TRAUMATIC ABDOMINAL WALL RUPTURE. Br J Surg. 1964 Feb;51:153–4. doi: 10.1002/bjs.1800510213. PMID: 14117773.

[15] Bertelsen S. The surgical treatment of spigelian hernia. Surg Gynecol Obstet. 1966 Mar;122(3):567–72. PMID: 5908668.

[16] Hurlbut HJ, Moseley T. Spigelian hernia in a child. South Med J. 1967 Jun;60(6):602 passim. doi: 10.1097/00007611-196706000-00009. PMID: 4225837.

[17] Graivier L, Alfieri AL. Bilateral Spigelian hernias in infancy. Am J Surg. 1970 Dec;120(6):817–9. doi: 10.1016/s0002-9610(70)80059-9. PMID: 4249783.

[18] Graivier L, Bernstein D, RuBane CF. Lateral ventral (spigelian) hernias in infants and children. Surgery. 1978 Mar;83(3):288–90. PMID: 24281.

[19] Graivier L, Bronsther B, Feins NR, Mestel AL. Pediatric lateral ventral (spigelian) hernias. South Med J. 1988 Mar;81(3):325–6. doi: 10.1097/00007611-198803000-00009. PMID: 3279528.

[20] Herbert RJ, Turner FW: Traumatic Abdominal Wall hernia in a 7-year-old child. J Pediatr Surg 12:609–610, 1977.

[21] Constantino L, Contestabile D, Rocua E: Strangulated spigelian hernia in a child Riv Chir Pediatr 16:236–238, 1974.

[22] Atiemo EA, Goswami G. Traumatic ventral hernia. J Trauma. 1974 Feb;14(2):181–2. doi: 10.1097/00005373-197402000-00011. PMID: 4272802.

[23] Houlihan TJ. A review of Spigelian hernias. Am J Surg. 1976 Jun;131(6):734–5. doi: 10.1016/0002-9610(76)90191-4. PMID: 937655.

[24] Jarvis PA, Seltzer MH. Pediatric Spigelian hernia: a case report. J Pediatr Surg. 1977 Aug;12(4):609–10. doi: 10.1016/0022-3468(77)90211-1. PMID: 894464.

[25] Bar-Maor, J.A., Sweed, Y. Spigelian hernia in children, two cases of unusual etiology. Pediatr Surg Int 4, 357–359 (1989). 10.1007/BF00183407

[26] Mitchiner JC. Handlebar hernia: diagnosis by abdominal computed tomography. Ann Emerg Med. 1990 Jul;19(7):812–3. doi: 10.1016/s0196-0644(05)81709-3. PMID: 2389864.

[27] Damschen DD, Landercasper J, Cogbill TH, Stolee RT. Acute traumatic abdominal hernia: case reports. J Trauma. 1994 Feb;36(2):273–6. doi: 10.1097/00005373-199402000-00026. PMID: 8114153.

[28] Kubalak G. Handlebar hernia: case report and review of the literature. J Trauma. 1994 Mar;36(3):438–9. PMID: 8145337.

[29] Komura J, Yano H, Uchida M, Shima I. Pediatric spigelian hernia: reports of three cases. Surg Today. 1994;24(12):1081–4. doi: 10.1007/BF01367460. PMID: 7780231.

[30] Pul N, Pul M. Spigelian hernia in children--report of two cases and review of the literature. Yonsei Med J. 1994 Mar;35(1):101–4. doi: 10.3349/ymj.1994.35.1.101. PMID: 8009893.

[31] Wright, J.E. Spigelian hernia in childhood. Pediatr Surg Int 9, 170–171 (1994). 10.1007/BF00179603

[32] Silberstein PA, Kern IB, Shi EC. Congenital spigelian hernia with cryptorchidism. J Pediatr Surg. 1996 Sep;31(9):1208–10. doi: 10.1016/s0022-3468(96)90233-x. PMID: 8887085.

[33] Iuchtman M, Kessel B, Kirshon M. Trauma-related acute spigelian hernia in a child. Pediatr Emerg Care. 1997 Dec;13(6):404–5. doi: 10.1097/00006565-199712000-00013. PMID: 9435002.

[34] Perez VM, McDonald AD, Ghani A, Bleacher JH. Handlebar hernia: a rare traumatic abdominal wall hernia. J Trauma. 1998 Mar;44(3):568. doi: 10.1097/00005373-199803000-00032. PMID: 9529196.

[35] Ostlie DJ, Zerella JT. Undescended testicle associated with spigelian hernia. J Pediatr Surg. 1998 Sep;33(9):1426–8. doi: 10.1016/s0022-3468(98)90027-6. PMID: 9766373.

[36] Kubota A, Shono J, Yonekura T, Hoki M, Asano S, Hirooka S, Kosumi T, Kato M, Oyanagi H. Handlebar hernia: case report and review of pediatric cases. Pediatr Surg Int. 1999 Jul;15(5-6):411–2. doi: 10.1007/s003830050616. PMID: 10415303.

[37] Al-Salem AH. Congenital spigelian hernia and cryptorchidism: cause or coincidence? Pediatr Surg Int. 2000;16(5-6):433–6. doi: 10.1007/s003839900292. PMID: 10955584.

[38] White JJ. Concomitant Spigelian and inguinal hernias in a neonate. J Pediatr Surg. 2002 Apr;37(4):659–60. doi: 10.1053/jpsu.2002.31635. PMID: 11912531.

[39] Losanoff JE, Richman BW, Jones JW. Spigelian hernia in a child: case report and review of the literature. Hernia. 2002 Dec;6(4):191–3. doi: 10.1007/s10029-002-0080-2. Epub 2002 Sep 7. PMID: 12424600.

[40] Fraser N, Milligan S, Arthur RJ, Crabbe DC. Handlebar hernia masquerading as an inguinal haematoma. Hernia. 2002 Mar;6(1):39–41. doi: 10.1007/s10029-002-0051-7. PMID: 12090581.

[41] Levy G, Nagar H, Blachar A, Ben-Sira L, Kessler A. Pre-operative sonographic diagnosis of incarcerated neonatal Spigelian hernia containing the testis. Pediatr Radiol. 2003 Jun;33(6):407–9. doi: 10.1007/s00247-003-0879-8. Epub 2003 Apr 12. PMID: 12692696.

[42] Goliath J, Mittal V, McDonough J. Traumatic handlebar hernia: a rare abdominal wall hernia. J Pediatr Surg. 2004 Oct;39(10):e20–2. doi: 10.1016/j.jpedsurg.2004.06.039. PMID: 15486881.

[43] Raveenthiran V. Congenital Spigelian hernia with cryptorchidism: probably a new syndrome. Hernia. 2005 Dec;9(4):378–80. doi: 10.1007/s10029-005-0316-z. Epub 2005 Mar 22. PMID: 15782280.

[44] Vaos G, Gardikis S, Zavras N. Strangulated low Spigelian hernia in children: report of two cases. Pediatr Surg Int. 2005 Sep;21(9):736–8. doi: 10.1007/s00383-005-1467-9. Epub 2005 Oct 20. PMID: 15977015.

[45] Torres de Aguirre A, Cabello Laureano R, García Valles C, Garrido Morales M, García Merino F, Martínez Caro A. Hernia de Spiegel: a propósito de 2 casos asociados a criptorquidia [Spigelian hernia: two cases associated to cryptorchidism]. Cir Pediatr. 2005 Apr;18(2):99–100. Spanish. PMID: 16044648.

[46] Durham MM, Ricketts RR. Congenital spigelian hernias and cryptorchidism. J Pediatr Surg. 2006 Nov;41(11):1814–7. doi: 10.1016/j.jpedsurg.2006.06.043. PMID: 17101349.

[47] O’Sullivan O, Bannon C, Clyne O, Flood H. Hypospadias associated undescended testis in a Spigelian hernia. Ir J Med Sci. 2006 Jan-Mar;175(1):77–8. doi: 10.1007/BF03169009. PMID: 16615238.

[48] Kumar, V. R. Ravi; Singal, Arbinder Kumar1,. Undescended testis in spigelian hernia. Journal of Indian Association of Pediatric Surgeons 12(4):p 233–234, Oct–Dec 2007. | DOI: 10.4103/0971-9261.40845

[49] Aksu B, Temizöz O, Inan M, Gençhellaç H, Başaran UN. Bilateral spigelian hernia concomitant with multiple skeletal anomalies and fibular aplasia in a child. Eur J Pediatr Surg. 2008 Jun;18(3):205–8. doi: 10.1055/s-2007-989486. PMID: 18493901.

[50] Litton K, Izzidien AY, Hussien O, Vali A. Conservative management of a traumatic abdominal wall hernia after a bicycle handlebar injury (case report and literature review). J Pediatr Surg. 2008 Apr;43(4):e31–2. doi: 10.1016/j.jpedsurg.2007.12.059. PMID: 18405697.

[51] Fascetti-Leon F, Gobbi D, Gamba P, Cecchetto G. Neonatal bilateral spigelian hernia associated with undescended testes and scalp aplasia cutis. Eur J Pediatr Surg. 2010 Mar;20(2):123–5. doi: 10.1055/s-0029-1220674. Epub 2009 Jun 9. PMID: 19513970.

[52] Christianakis, E., Paschalidis, N., Filippou, G. et al. Low Spigelian hernia in a 6-year-old boy presenting as an incarcerated inguinal hernia: a case report. J Med Case Reports 3, 34 (2009). 10.1186/1752-1947-3-34

[53] Rushfeldt C, Oltmanns G, Vonen B. Spigelian-cryptorchidism syndrome: a case report and discussion of the basic elements in a possibly new congenital syndrome. Pediatr Surg Int. 2010 Sep;26(9):939–42. doi: 10.1007/s00383-010-2681-7. Epub 2010 Aug 3. PMID: 20680633; PMCID: PMC2923717.

[54] Vega Y, Zequeira J, Delgado A, Lugo-Vicente H. Spigelian hernia in children: case report and literature review. Bol Asoc Med P R. 2010 Oct-Dec;102(4):62–4. PMID: 21766551.

[55] Singal AK, Ravikumar VR, Kadam V, Jain V. Undescended testis in Spigelian hernia--a report of 2 cases and review of the literature. Eur J Pediatr Surg. 2011 May;21(3):194–6. doi: 10.1055/s-0031-1271634. Epub 2011 Feb 18. PMID: 21337299.

[56] Inan M, Basaran UN, Aksu B, Dortdogan Z, Dereli M. Congenital Spigelian hernia associated with undescended testis. World J Pediatr. 2012 May;8(2):185–7. doi: 10.1007/s12519-011-0313-5. Epub 2011 Aug 27. PMID: 21874609.

[57] Lopez R, King S, Maoate K, Beasley S. Trauma may cause Spigelian herniae in children. ANZ J Surg. 2010 Sep;80(9):663. doi: 10.1111/j.1445-2197.2010.05414.x. PMID: 20840415.

[58] Yan J, Wood J, Bevan C, Cheng W, Wilson G. Traumatic abdominal wall hernia--a case report and literature review. J Pediatr Surg. 2011 Aug;46(8):1642–5. doi: 10.1016/j.jpedsurg.2011.04.004. PMID: 21843736.

[59] Lopez R, King S, Maoate K, Beasley S. Laparoscopic repair of paediatric traumatic Spigelian hernia avoids the need for mesh. ANZ J Surg. 2011 May;81(5):396–7. doi: 10.1111/j.1445-2197.2011.05720.x. PMID: 21518204.

[60] Bilici S, Güneş M, Göksu M, Melek M, Pirinçci N. Undescended testis accompanying congenital Spigelian hernia: is it a reason, a result, or a new syndrome? Eur J Pediatr Surg. 2012 Apr;22(2):157–61. doi: 10.1055/s-0032-1308702. Epub 2012 Apr 19. PMID: 22517524.

[61] Rathore A, Simpson BJ, Diefenbach KA. Traumatic abdominal wall hernias: an emerging trend in handlebar injuries. J Pediatr Surg. 2012 Jul;47(7):1410–3. doi: 10.1016/j.jpedsurg.2012.02.003. PMID: 22813805.

[62] Decker S, Engelmann C, Krettek C, Müller CW. Traumatic abdominal wall hernia after blunt abdominal trauma caused by a handlebar in children: a well-visualized case report. Surgery. 2012 Jun;151(6):899–900. doi: 10.1016/j.surg.2011.02.020. Epub 2011 May 5. PMID: 21549405.

[63] Parihar D, Kadian YS, Raikwar P, Rattan KN. Congenital spigelian hernia and cryptorchidism: another case of new syndrome. APSP J Case Rep. 2013 Oct 23;4(3):41. PMID: 24381837; PMCID: PMC3863830.

[64] Thakur SK, Gupta S, Goel S. Traumatic spigelian hernia due to handlebar injury in a child: a case report and review of literature. Indian J Surg. 2013 Jun;75(Suppl 1):404–6. doi: 10.1007/s12262-012-0734-y. Epub 2012 Sep 18. PMID: 24426630; PMCID: PMC3693233.

[65] Upasani A, Bouhadiba N. Paediatric abdominal wall hernia following handlebar injury: should we diagnose more and operate less? BMJ Case Rep. 2013 Apr 19;2013:bcr2012008501. doi: 10.1136/bcr-2012-008501. PMID: 23606382; PMCID: PMC3644890.

[66] Balsara ZR, Martin AE, Wiener JS, Routh JC, Ross SS. Congenital spigelian hernia and ipsilateral cryptorchidism: raising awareness among urologists. Urology. 2014 Feb;83(2):457–9. doi: 10.1016/j.urology.2013.09.032. Epub 2013 Nov 26. PMID: 24286599.

[67] Spinelli C, Strambi S, Pucci V, Liserre J, Spinelli G, Palombo C. Spigelian hernia in a 14-year-old girl: a case report and review of the literature. European J Pediatr Surg Rep. 2014 Jun;2(1):58–62. doi: 10.1055/s-0034-1370771. Epub 2014 Mar 12. PMID: 25755973; PMCID: PMC4336063.

[68] Talutis SD, Muensterer OJ, Pandya S, McBride W, Stringel G. Laparoscopic-assisted management of traumatic abdominal wall hernias in children: case series and a review of the literature. J Pediatr Surg. 2015 Mar;50(3):456–61. doi: 10.1016/j.jpedsurg.2014.10.020. Epub 2014 Dec 9. PMID: 25746707.

[69] Pederiva F, Guida E, Maschio M, Rigamonti W, Gregori M, Codrich D. Handlebar injury in children: The hidden danger. Surgery. 2016 May;159(5):1477. doi: 10.1016/j.surg.2015.08.009. Epub 2015 Sep 19. PMID: 26387787.

[70] Montalvo Ávalos C, Álvarez Muñoz V, Fernández García L, López López AJ, Oviedo Gutiérrez M, Lara Cárdenas C, et al. Atypical hernia defects in childhood. Rev Pediatr Aten Primaria. 2015;17:139–43.

[71] Volpe A, Virgone C, Gamba P. Successful conservative management of handlebar hernia in children. Pediatr Int. 2017 Jan;59(1):105–106. doi: 10.1111/ped.13110. PMID: 28102633.

[72] Shea B, Fasano G, Cohen IT, Pediatric Spigelian hernia: A case report and review of the literature, Journal of Pediatric Surgery Case Reports (2017), doi: 10.1016/j.epsc.2017.01.014.

[73] Kamal N R, Shubhangi S. Spigelian Hernia in 2-Year-Old Male Child: A Rare Case Report. JOJ Case Stud. 2017; 3(3) : 555612. DOI: 10.19080/JOJCS.2017.03.555612

[74] Rinaldi VE, Bertozzi M, Magrini E, Riccioni S, Di Cara G, Appignani A. Traumatic Abdominal Wall Hernia in Children by Handlebar Injury: When to Suspect, Scan, and Call the Surgeon. Pediatr Emerg Care. 2020 Sep;36(9):e534–e537. doi: 10.1097/PEC.0000000000001153. PMID: 28441239.

[75] Sinopidis X, Panagidis A, Alexopoulos V, Karatza A, Georgiou G. Congenital Spigelian Hernia Combined with Bilateral Inguinal Hernias. Balkan Med J. 2018 Sep 21;35(5):402–403. doi: 10.4274/balkanmedj.2017.1306. Epub 2018 Apr 4. PMID: 29636314; PMCID: PMC6158471.

[76] Sengar M, Mohta A, Neogi S, Gupta A, Viswanathan V. Spigelian hernia in children: low versus classical. J Pediatr Surg. 2018 Nov;53(11):2346–2348. doi: 10.1016/j.jpedsurg.2018.06.016. Epub 2018 Jun 23. PMID: 30017065.

[77] So HF, Nabi H. Handlebar hernia - A rare complication from blunt trauma. Int J Surg Case Rep. 2018;49:118–120. doi: 10.1016/j.ijscr.2018.06.003. Epub 2018 Jun 19. PMID: 30005362; PMCID: PMC6037005.

[78] Vega-Mata N, Vázquez-Estevez JJ, Montalvo-Ávalos C, Raposo-Rodríguez L. Laparoscopic Spigelian hernia repair in childhood. Literature review [Abordaje laparoscópico de una hernia de Spiegel en edad pediátrica. Revisión de la literatura]. Cir Cir. 2019;87(1):101–105. Spanish. doi: 10.24875/CIRU.18000338. PMID: 30600802.

[79] Deshmukh SS, Kothari PR, Gupta AR, Dikshit VB, Patil P, Kekre GA, Deshpande A, Kulkarni AA, Hukeri A. Total laparoscopic repair of Spigelian hernia with undescended testis. J Minim Access Surg. 2019 Jul-Sep;15(3):265–267. doi: 10.4103/jmas.JMAS_196_18. PMID: 30618422; PMCID: PMC6561065.

[80] Nagara S, Fukaya S, Muramatsu Y, Kaname T, Tanaka T. A case report of rare ZC4H2-associated disorders associated with three large hernias. Pediatr Int. 2020 Aug;62(8):985–986. doi: 10.1111/ped.14211. Epub 2020 Jul 20. PMID: 32686882.

[81] Taha A, Algethami NE, AlQurashi R, Alnemari AK. Outcome of Orchidopexy in Spigelian Hernia-Undescended Testis Syndrome. Cureus. 2021 Mar 5;13(3):e13714. doi: 10.7759/cureus.13714. PMID: 33833925; PMCID: PMC8018874.

[82] García Sánchez P, Bote Gascón P, González Bertolín I, Bueno Barriocanal M, López López R, de Ceano-Vivas la Calle M. Handlebar Hernia: An Uncommon Traumatic Abdominal Hernia. Pediatr Emerg Care. 2021 Dec 1;37(12):e879–e881. doi: 10.1097/PEC.0000000000002267. PMID: 33105464.

[83] Sinacer S, Semari BZ, Khemari S, Kharchi A, Haif A, Soualili Z. Congenital Spigelian hernia in a neonate associated with several anomalies: A case report. J Neonatal Surg [Internet]. 2021Aug.13 [cited 2024May5];10:38. Available from: https://www.jneonatalsurg.com/ojs/index.php/jns/article/view/972

[84] Thamri F, Houidi S, Zouaoui A, et al. Traumatic Spigelian hernia in a child. J Pediatr Surg Case Rep. 2021;75:102099.

[85] Gonuguntla A, Thotan SP, Pai N, Kumar V, Prabhu SP. Congenital Spigelian Hernia With Ipsilateral Ectopic Testis. Ochsner J. 2022 Fall;22(3):277–280. doi: 10.31486/toj.21.0134. PMID: 36189089; PMCID: PMC9477124.

[86] Kropilak AD, Sawaya DE. Traumatic Spigelian Hernia in a Pediatric Patient Following a Bicycle Injury. Am Surg. 2022 Aug;88(8):1933–1935. doi: 10.1177/00031348221087354. Epub 2022 Apr 7. PMID: 35389281.

[87] Okumuş M, Zerbaliyev E, Akdağ A. Congenital spigelian hernia and ipsilateral undescended testis: An ongoing etiological debate – A case report. Int J Abdom Wall Hernia Surg 2022;5:209–11.

[88] Kangabam B. Traumatic Spigelian Hernia Following Blunt Abdominal Trauma. Cureus. 2023 Feb 27;15(2):e35564. doi: 10.7759/cureus.35564. PMID: 37007403; PMCID: PMC10063246.

[89] Farina R, Pennisi M, Desiderio C, Valerio Foti P, D’Urso M, Inì C, Motta C, Galioto S, Garofalo A, Clemenza M, Ilardi A, Lavalle S, Basile A. Spigelian-cryptorchidism syndrome: Lesson based on a case report. Radiol Case Rep. 2024 May 23;19(8):3372–3375. doi: 10.1016/j.radcr.2024.04.080. PMID: 38827042; PMCID: PMC11143774.

[90] Ablatt S, Escobar MA Jr. Spigelian hernia and cryptorchidism syndrome: open spigelian hernia repair and laparoscopic one-stage orchiopexy for ectopic testis. BMJ Case Rep. 2025 Jan 4;18(1):e261858. doi: 10.1136/bcr-2024-261858. PMID: 39755551; PMCID: PMC11751608.

