## Supplementary material for "Pediatric Spigelian Hernia and Spigelian-Cryptorchidism Syndrome: an Integrating Review": Table 1..

| **Author** | **Country** | **Patient´s age** | **Patient´s Sex** | **Laterality** | **Associated malformations/anomalies** | **Complications associated with SP** | **Relevant**  **background** | **Defect size** | **Radiological studies** | **Treatment** | **Surgical approach/findings** | **Surgical outcome** |
| --- | --- | --- | --- | --- | --- | --- | --- | --- | --- | --- | --- | --- |
| Scopinaro et al. (1935) [9] | - | 6-days-old | Male | - | - | Strangulation | - | - | - | - | - | Death |
| Hurwitt et al. (1955) [10] | - | 8y | Male | Bilateral | - | - | - | Right side: 2,6 cm | - | - | - | - |
| Landry et al. (1956) [11] | - | 14y | Male | Left | - | - | Previous local trauma | 4cm | - | - | - | - |
| Isaacson et al. (1956) [12]* | USA | 3y | Male | Right | - | - | Nail puncture | - | - | - | - | - |
| Wilson et al. (1959) [13] | USA | 2.5y | Male | Left | - | - | Previous local trauma (automobile) | - | Plain abdominal x-ray: traumatic hernia of the abdominal wall | Surgical | ESR 6 days after the accident  Hernial sac containing omentum and transverse colon.  Repair: Interrupted silk suture | - |
| Roberts et al. (1964) [14]***** | UK | 9y | Male | Left | - | - | Previous local trauma (bicycle handlebar) | 7x3 cm | - | Surgical | Repair: Interrupted catgut suture | Favorable (6m follow-up) |
| Bertelsen S (1966) [15]* | - | 9y | Male | Right | - | - | - | 2x2 cm | - | - | - | Favorable |
| Hurlbut et al. (1967) [16] | USA | 8y | Male | Right | - | - | Previous local trauma | 3x5 cm | Plain abdominal X-ray: gas-filled small bowel loops compatible with incarcerated hernia | Surgical | ESR | Favorable |
| Graivier et al. (1967-1988) [17-19]. | USA | 1) 10m  2) 6m  3) 6m  4) 9m  5) 15y  6)17y  7)15m  8)4y  9)10d | 1) Male  2) Male  3) Male  4) Male  5) Female  6) Female  7) Female  8) Female  9) Female | 1) Bilateral  2) Right  3) Right  4) Left  5) Right  6) Bilateral  7) Bilateral  8) Left  9) Left | 1) Umbilical Hernia  and bilateral indirect  Inguinal hernias | - | - | 1) 2x5 cm (left)  1x3 cm (right) | - | Surgical | Patients 1-8: ESR  Patient 9: loss of follow-up previous to surgery  Hernial sacs: resected or reduced  Repair: Interrupted silk suture | Favorable. No recurrences  (follow-up up to 19y) |
| Herbert et al. (1973) [20] | Canada | 7y | Male | Left | - | - | Previous local trauma (bicycle handlebar) | 4cm | Plain Abdominal X-ray: acute gastric dilatation | Surgical | ESR 3 weeks after the accident. SH Repair: Interrupted 2-0 polyglycolic acid suture. | Favorable |
| Constantino et al. (1974) [21] | - | 8y | Male | Left | - | Strangulation | - | - | - | - | - | Favorable |
| Atiemo et al. (1974) [22]***** | Ghana | 6y | Male | Left | Splenic enlargement (physical examination) | - | Previous local trauma (cow goring) | - | - | Surgical | ESR 6 days after the accident. Full abdominal wall hernia. Primary repair. | Favorable |
| Houlihan et al. (1976) [23] | USA | 21y | Male | Bilateral | - | - | - | - | - | - | ESR | Favorable |
| Jarvis et al. (1977) [24] | USA | 13y | Female | Left | - | - | - | 3 cm | - | Surgical | SH. Peritoneal sac present. | - |
| Bar-Maor et al. (1989) [25] | Israel | 1) 5y  2) 3m | 1) Female  2) Male | 1) Right  2) Left | 2) Left Bochdalek hernia | - | 1) Abdominal blunt trauma (road accident)  2) Previous abdominal surgery with forceful stretching of the abdominal wall (Bochdalek hernia) | 1) 4 cm  2) 1 cm | - | 1) Conservative (spontaneous resolution)  2) Surgical | ESR. SH | Favorable |
| Mitchiner et al. (1990) [26]***** | USA | 7y | Male | Left | - | Incarceration | Previous local trauma (bicycle handlebar) | 6 cm | CT (oral and intravenous contrast media): rent in the anterior abdominal wall with a loop of small bowel exteriorized in the subcutaneous tissue | Surgical  (Urgent) | 3 feet of viable small bowel found in the subcutaneous tissue  protruding through a large fascial defect. Reduction and primary repair. | Favorable (4 months follow-up) |
| Damschen et al. (1994) [27]***** | USA | 5y | Male | Right | - | - | Previous local trauma (bicycle handlebar) | - | - | Surgical | Urgent surgery: SH. Small bowel herniating through the muscle layers. Repair: Interrupted polyglycolic acid suture. | Lost to follow-up (unknown) |
| Kubalak G (1994) [28]***** | USA | 8y | Male | Right | - | - | Previous local trauma (bicycle handlebar) | 6 cm | - | - | - | Favorable (2y follow-up) |
| Komura et al. (1994) [29] | Japan | 1) 6m  2) 8m  3) 3y | 1) Female  2) Female  3) Male | 1) Right  2) Right  3) Left | 1) Ipsilateral mediastinal neuroblastoma (10-12th IC spaces) - Incidental diagnosis of tumor during ultrasound examination of the SH.  2) Ipsilateral mediastinal neuroblastoma (9-10th IC spaces) | - | 2) Previous tumor extirpation (1 week before) | 1) 5x3 cm  2) 4cm  3) 7cm | 3) US: hernial orifice 4x3 cm. Thin muscular layer in the area (3 mm). MRI: fatty infiltration of internal oblique/transverse muscles | 1) Surgical  2) Conservative  3) Surgical | 1) SH. Peritoneal sac present. Primary repair (mattress 3-0 silk sutures)**  3) SH. Primary repair (mattress 2-0 silk sutures)** | 1) Favorable (5y follow-up)  2) Favorable (spontaneous resolution) ***  3) Favorable (2m follow-up) |
| Pul et al. (1994) [30] | Turkey | 1) 18m  2) 2,5m | 1) Male  2) Male | 1) Right  2) Right | 1) Right UDT  2) Right indirect inguinal hernia | - | - | 1) 7x7 cm  2) 4x4 cm |  | 1) Surgical | 1) SH. Small ring defect (1x1cm). Peritoneal sac present and excised. Primary repair.  2) SH. Small ring defect (2 cm). Peritoneal sac present and excised. Primary repair. | 1) Favorable  2) Favorable |
| Wright JE. (1994) [31] | Australia | 1) 19m  2) 5y  3) 7y | 1) Male  2) Male  3) Male | 1) Right  2) Right  3) Left | - | - | - | - | - | 3) Surgical | 3) SH. Peritoneal sac present (not excised). Two-layer repair | 3) Favorable (8y follow-up) |
| Silberstein et al. (1996) [32] | Australia | 1) Newborn  2) Newborn | 1) Male  2) Male | 1) Left  2) Right | 1) Contralateral generalized abdominal musculature weakness. Left UDT.  2) Right UDT. | - | - | 2) 3 cm | - | 1) Surgical (10w of age)  2) Surgical (4.5m of age) | 1) SH. Peritoneal sac present containing left testis (small in size, without testicular-epididymal dissociation). Absence of inguinal canal. Primary repair and orchidopexy.  2) HS. The internal oblique  and transversalis muscles were poorly developed. Peritoneal sac present containing right testis. Absence of inguinal canal. Primary repair and orchidopexy. | - |
| Iuchtman et al. (1997) [33] | Israel | 7y | Male | - | - | - | Previous local trauma (bicycle) | - | US: normal | Surgical | Urgent surgery: SH. Peritoneal protrusion (intact peritoneum). Primary repair without mesh. | Favorable |
| Pérez et al. (1998) [34]***** | USA | 11y | Male | Left | - | - | Previous local trauma (bicycle handlebar) | - | - | Surgical | Urgent surgery: SH. Peritoneal tearing. Repair in layers. | Favorable |
| Ostlie et al. (1998) [35] | USA | Newborn | Male | Right | 1) Right UDT. The right testis was palpable on the hernial sac. | - | - | 1.5 cm | - | Surgical | SH. Thinning of the layers. Absence of inguinal canal. Peritoneal sac containing the right testis. Primary repair (absorbable sutures) and orchidopexy. | - |
| Kubota et al. (1999) [36]***** | Japan | 9y | Male | Right | - | - | Previous local trauma (bicycle handlebar) | 4 cm | CT: rent in the abdominal wall through which  intestinal loops protruded into the subcutaneous space. | Surgical | SH. Peritoneal rupture. Primary repair | Favorable |
| Al-Salem et al. (2000) [37] | Saudi Arabia | 1) 3m  2) Newborn | 1) Male  2) Male | 1) Left  2) Left | 1) Left UDT.  2) Micrognathia, cleft palate, malformed ears, right clubfoot, malformed left lower limb, left UDT. Left testis palpable on the hernial sac. (Insulin-dependent diabetic mother). | - | - | 1) 5 cm | 1) US: normal | 1) Surgical | 1) SH. Peritoneal sac present containing left testis (small in size, without testicular-epididymal dissociation) and sigmoid colon. Primary repair and orchidopexy. | 1) Favorable (2.5y follow-up)  2) Died before surgery (sepsis, not related to SH) |
| White J (2002) [38] | USA | 1m | Female | Right | Bilateral inguinal hernia. | Incarceration/small bowel obstruction. | SH appeared in the immediate postoperative period after bilateral inguinal hernia repair. | 1.5 cm (ring) | Plain abdominal X-ray: Partial small bowel obstruction  US: bowel in the mass, under the skin | Surgical | SH. Reduction of the small bowel, primary repair. | Favorable |
| Losanoff et al. (2002) [39] | USA | 12y | Male | Right | - | Omentum incarceration mimicking acute appendicitis. | . | 1.5 cm | Plain chest and abdominal X-rays: Normal | Surgical (urgent) | Normal appendix. SH with incarcerated and infarcted omentum. Resection, primary repair | Favorable (6m follow-up) |
| Fraser et al. (2002) [40]***** | UK | 11y | Male | Right | - | Inguinal hematoma | Previous local trauma (bicycle handlebar) | 12x8 cm (bulge) | US: disruption of the muscle layers of the  abdominal wall with bowel and free fluid lateral to the rectus  muscle.  Plain abdominal X-ray (lateral): A collection of gas was visible in the anterior abdominal | Surgical | Traumatic hernia involving all layers of the lower abdominal wall above the inguinal canal. Primary repair (absorbable suture). | Favorable |
| Levy et al. (2003) [41] | Israel | 1) 1m  2) 5w | 1) Male  2) Male | 1) Bilateral  2) Left | 1) Bilateral UDT.  2) Left UDT. | 1) Incarceration | - | 2) 5 cm | 1) US (Right side): HS containing Incarcerated bowel loops and the undescended right testis  US (Left side): small SH containing the undescended left testis  2) US: left SH containing bowel loops and the undescended left testis | 1) Right side: Surgical (urgent)  Left side:  Surgical (deferred, with 10m)  2) Surgical (deferred, with 2m) | 1) Right side: SH. Small bowel reduction, primary repair, right orchidopexy  Left side: SH primary repair, left orchidopexy  2) SH. Peritoneal sac present containing small bowel and left testis. Reduction, primary repair, and orchidopexy | Favorable |
| Goliath et al. (2004) [42] ***** | USA | 11y | Male | Right | - | - | Previous local trauma (bicycle handlebar) | - | CT: Intestinal  loops protruding through a defect in  the abdominal wall into the subcutaneous  space. | Surgical | Defect throughout his entire abdominal  wall, including the fascia, muscular layers, and peritoneum, with bowel  protruding into the subcutaneous space, leaving his skin and intraabdominal  organs completely intact. Primary repair | Favorable |
| Raveenthiran V (2005) [43] | India | Newborn (2d) | Male | Right | Imperforate anus. Bilateral UDT. Hypoplastic scrotum. Left inguinal hernia (noted with 1m of life), umbilical hernia (noted with 2m of life) | - | - | 5x5cm | US: bowel loops in the intermuscular plane at the site of the SH | Surgical (deferred, with 13m) | SH. Peritoneal sac present containing right testis. Absence of inguinal canal. | Favorable |
| Vaos et al. (2005) [44] | Greece | 1) 20m  2) 8m | 1) Male  2) Male | 1) Left  2) Right | - | 1) Strangulation  2) Strangulation | - | 1) 2.5 cm (bulge), 2 cm (ring)  2) 2 cm (bulge), 1 cm (ring) | 1) Plain abdominal X-ray: several gas-fluid levels in the small bowel | 1) Surgical (urgent)  2) Surgical (urgent) | 1) SH. Peritoneal sac present containing incarcerated viable small bowel loops and infarcted omentum. Omentectomy, reduction, primary repair.  2) SH. Peritoneal sac present containing incarcerated viable small bowel loops. Reduction, primary repair. | 1) Favorable (12m follow-up)  2) Favorable (6m follow-up) |
| Torres de Aguirre (2005) [45] | Spain | 1) Newborn  2) Infant (5w) | 1) Male  2) Male | 1) Right  2) Bilateral | 1) Right UDT.  2) Bilateral UDT. | 1) Incarceration  2) Right incarceration | - | - | 1) US and Plain abdominal x-ray: incarcerated right inguinal hernia | 1) Surgical (urgent)  2) Surgical (urgent) | 1) SH. Peritoneal sac present containing incarcerated small bowel and right testis. Reduction, primary repair, and orchidopexy  2) Bilateral SH. Right: Peritoneal sac present containing incarcerated viable small bowel loops. Reduction, primary repair. Left: peritoneal sac present containing a hypoplastic left testis with epididymal-testicular dissociation. Orchiectomy and primary repair. | Favorable |
| Durham et al. (2006) [46] | USA | 1) 8m  2) 13m  3) 14m****  4) 2m**** | 1) Male  2) Male  3) Male  4) Male | 1) Left  2) Bilateral  3) Bilateral  4) Right | 1) Left UDT  2) Bilateral UDT  3) Bilateral UDT  4) Bilateral UDT | - | - | - | - | Surgical | 1) SH. Peritoneal sac present containing left testis. Orchidopexy and primary repair  2) bilateral SH. Staged repair: right side with 13m and left with 16m. Peritoneal sac present containing testes. Orchidopexy and primary repair (absorbable sutures, 8-ply SIS mesh on right side)  3) bilateral SH. Repair. Testicles found in their ipsilateral SH sac. Orchidopexy and primary repair  4) With 13m: Right intraabdominal testis: orchidopexy. Right SH: primary repair. With 16m: left inguinal orchidopexy. | 1) Complication: scrotal abscess. Loss of follow-up  2) Favorable (laxity on the right side where the SIS patch was used)  3) Favorable (45m follow-up)  4) Favorable (12m follow-up) |
| O´Sullivan et al. (2006) [47] | Ireland | 4m | Male | Left | Left UDT. Hypospadias. | - | - | - | - | Surgical | SH. Peritoneal sac present containing left testis. Orchidopexy, primary repair. | - |
| Kumar et al. (2007) [48] | India | 3y | Male | Right | Right UDT. | - | - | 7 cm (neck of peritoneal sac) | - | Surgical | SH. Peritoneal sac present containing right testis. Orchidopexy, primary repair (VYPRO mesh) | Favorable |
| Aksu et al. (2007) [49] | Turkey | 4y | Female | Bilateral | Consanguineous parents. Multiple skeletal anomalies. | - | - | 6.5 cm (bulges),  3x2 cm (right ring)  2x1 cm (left ring) | - | Surgical | Bilateral SH. Internal oblique and transverse abdominus missing. Peritoneal sac present containing small bowel loops. Reduction, primary repair (3/0 Vicryl sutures). | Favorable (8m follow-up) |
| Litton et al. (2008) [50]***** | UK | 13y | Male | Right | - | - | Previous local trauma (bicycle handlebar) |  | CT: herniation of bowels. No intraabdominal injury | Conservative (spontaneous resolution) | - | Favorable (4m follow-up) |
| Fascetti-Leon et al. (2009) [51] | Italy | Newborn | Male | Bilateral | Bilateral UDT. Scalp aplasia cutis. Hypertelorism, delayed growth, small head circumference, hypoplasia of the nasal alae, dentition anomalies | - | - | - | US: bilateral SH. Bilateral UDT | Surgical | Bilateral SH. Peritoneal sac present containing testes. Orchidopexy and primary repair (VICRYL^TM^ mesh) | Favorable (12m follow-up) |
| Christianakis et al. (2009) [52] | Greece | 6y | Male | Left | - | Inguinoscrotal pain | - | 1.5 cm (ring) | - | Surgical | SH. Peritoneal sac present. Primary repair (non-absorbable sutures) | Favorable (8y follow-up) |
| Rushfeldt et al. (2010) [53] | Norway | Newborn | Male | Right | Right UDT | Incarceration | - | - | US: SH with hernial sac between the obliquus externus and the obliquus internus (7mm hernia opening) containing right testis and a loop of small bowel. | Surgical (urgent) | SH. Peritoneal sac present containing incarcerated viable small bowel loops and right testis Reduction, orchidopexy, primary repair. | Favorable |
| Vega et al. (2010) [54] | Puerto Rico | 9y | Male | Left | - | - | - | - | - | Surgical | SH. Primary repair. | - |
| Singal et al. (2011) [55] | India | 1) 3y  2) 3m | 1) Male  2) Male | 1) Right  2) Left | 1) Right UDT  2) Left UDT. Glanular hypospadias | - | - | 1) 7 cm (ring/neck) | - | Surgical | 1) SH. Thinned out internal oblique. Peritoneal sac present containing right testis. Orchidopexy  and primary repair (VYPRO mesh)  2) SH. Peritoneal sac present containing left testis. No inguinal canal nor gubernaculum present. Orchidopexy and primary repair | 1) Favorable (4y follow-up)  2) Favorable (1y follow-up) |
| Inan et al. (2011) [56] | Turkey | Newborn (26d) | Male | Right | Right UDT | - | - | 3 cm (bulge), 2 cm (ring) | US: fascial plane defect through the linea  semilunaris with herniation of bowel loops between the  internal and external oblique muscles  . Absence of testis in right scrotum. No inguinal canal nor spermatic cord. | Surgical | SH. Thinning of layers. Peritoneal sac present containing right testis. Orchidopexy and primary repair. | Complication: scrotal abscess (postoperative day 8) and atrophy of the testis |
| Lopez et al. (2011) [57,58] | New Zealand | 14y | Male | Left | - | - | Previous local trauma (bicycle handlebar) | - | CT: herniation of fat and vessels  through a defect in the abdominal wall musculature consistent with  a diagnosis of SH | Surgical | SH. Laparoscopic repair | - |
| Yan et al. (2011) [59] ***** | Australia | 8y | Male | Right | - | - | Previous local trauma (BMX handlebar) | - | CT: TAWH | Surgical (urgent) | TAWH. Primary repair | Favorable (1m follow-up) |
| Bilici et al. (2012) [60] | Turkey | 1) 6m  2) 1y  3) 2y  4) 5y | 1) Male  2) Male  3) Male  4) Male | 2 patients: left  2 patients: right | 4 patients: Ipsilateral UDT | 4 patients: abdominal distension | - | 1.5 to 2.5 cm | - | Surgical | SH. Peritoneal sac present containing ipsilateral testis (all cases). Absence of inguinal canal Orchidopexy and primary repair | Favorable (6m follow-up) |
| Rathore et al. (2012) [61] ***** | USA | Five patients (9-15y)  1) 15y  2) 15y  3) 13y  4) 9y  5) 11y | All male | - | - | Three patients with associated visceral injury: 1,4) cecal wall hematoma, 2) duodenal hematoma, pancreatic contusion | Previous local trauma (bicycle handlebar) in all patients | - | CT (patient no.5): compatible with an SH | Surgical | - | 2) pancreatic pseudocyst. 1) persistent pain. Three cases evolved favorably. |
| Decker et al. (2012) [62] ***** | Germany | 13y | Male | Right | - | Abdominal wall hematoma | Previous local trauma (bicycle handlebar) | - | CT: 18.39 mm gap in the fascia of the abdominal rectus muscle and the internal and external oblique muscles with two intestinal loops | Surgical | SH. Primary repair. | Favorable |
| Parihar et al. (2013) [63] | India | 3m | Male | Right | Right UDT | - | - | - | US: testis in the layers of the abdominal wall, along with small bowel loops, echoes. | Surgical | SH. Peritoneal sac present containing ipsilateral testis. Absence of inguinal canal. Orchidopexy and primary repair | Favorable |
| Thakur et al. (2013) [64] | India | 9y | Male | Right | - | - | Previous local trauma (bicycle handlebar) 5 weeks before | 6x4 cm (bulge),  4x4 cm (ring) | US: 4x4 cm defect along the right semilunar line with small bowel loops | Surgical | SH. Peritoneal sac present (opened). Primary repair (non-absorbable sutures) | Favorable (18m follow-up) |
| Upasani et al. (2013) [65] ***** | UK | 12y | Male | Left (upper) | - | - | Previous local trauma (BMX handlebar) | 10x10 cm (bulge),2cm (ring) | US: abdominal wall hematoma. CT: 2 cm fascial defect with fat herniating through the defect | Conservative (partial resolution) | - | Favorable (6m follow-up) |
| Balsara et al. (2014) [66] | USA | Newborn (2w) | Male | Left | Left UDT | - | - | - | US: normal-sized left testicle within the SH in the left lower quadrant with loops of bowel. | Surgical | Exploratory laparoscopy: vas deferens and spermatic vessels entering the hernia sac. Left testis confirmed to lie within the sac itself. Open correction: SH. Hernial sac present containing left testis. Orchidopexy and primary repair. | Favorable (7m follow-up) |
| Spinelli et al. (2014) [67] | Italy | 14y | Female | Right | - | One-year history of recurrent abdominal pain. | - | 1.5 cm | US: fascial defect in right hemiabdomen | Surgical | SH. Hernia lipoma. Peritoneal sac present containing greater omentum. Reduction and primary repair. | Favorable |
| Talutis et al. (2014) [68] | USA | 1) 9y  2) 7y  3) 11y  4) 7y | 1) Male  2) Male  3) Male  4) Female | 1) Left  2) Right  3) Left  4) Right | - | 1) Contusion to the mid-jejunum.  2) Mesenteric defect in the ileocecal region  4) Ileal perforation | 1,3,4) Previous local trauma (bicycle handlebar)  2) ATV collision | 1) 4x3 cm | 1) CT: 4x3 cm fascial defect (SH) with a contusion to the mid-jejunum.  4) CT: Handlebar sign. Evidence of RLQ abdominal wall defect with herniation | 1-3) Surgical | 1,3,4) Laparoscopic. Conversion to open surgery.  2) Open surgery | 1-4) Favorable |
| Pederiva et al. (2015) [69] ***** | Italy | 9y | Male | Left | - | Ileal perforation (not diagnosed at CT) | Previous local trauma (bicycle handlebar) | - | CT: abdominal wall hematoma. Defect through rectus sheath between left rectus abdominis and internal and external oblique. Intraabdominal fat herniated through the defect. | Surgical | Laparotomy. Intestinal resection. Primary repair (absorbable sutures). | Favorable |
| Montalvo et al. (2015) [70] | Spain | 1m | Female | Left | Left UDT | - | - | - | US (NR) | - | SH. Laparoscopic approach: Peritoneal sac present containing left testis. Orchidopexy. NO primary repair | Favorable (unknown follow-up) |
| Volpe et al. (2016) [71] ***** | Italy | 1) 8y  2) 9y | 1) Male  2) Male | 1) Right  2) Right | - | - | 1) Previous local trauma (bicycle handlebar)  2) Previous local trauma (bicycle handlebar) | 1) 1.5 cm  2) 1 cm | 1) US: 1.5 cm hernia between the rectus and the internal oblique with herniation of the omentum  2) US: 1 cm defect with bowel herniation | 1) Conservative  2) Conservative | - | 1) Favorable (last control: 3mm defect) (12m follow-up  2) Favorable (last control: 3 mm defect) (2m follow-up |
| Shea et al. (2017) [72] | USA | 16y | Male | Left | - | - | Previous local trauma (bicycle handlebar) | 1x1cm | CT: SH in the anterior left lower abdominal wall. | Surgical | SH. No peritoneal sac. Primary repair (absorbable sutures) | Favorable |
| Kamal et al. (2017) [73] | India | 2y | Male | Right | Patchy frontal hair loss, left eye deviation, bilateral UDT. | - | - | - | US: defect in the anterior abdominal wall lateral to the rectus muscle with herniation of preperitoneal fat. Absence of both testes in the scrotal sac. CT: SH with small bowel herniation. Both testes on the inguinal region | Surgical | SH. Peritoneal sac present. Primary repair  One month later, bilateral orchidopexy. | Favorable |
| Rinaldi et al. (2017) [74] ***** | Italy | 1) 12y  2) 13y | 1) Male  2) Male | 1) Left  2) Right | - | - | 1) Previous local trauma (bicycle fall)  2) Previous local trauma (bicycle fall) | 1) 5 cm | 1) US: no peritoneal lesions; CT: defect in left anterior abdominal wall between lateral margin of the left rectus and medial margin of ipsilateral oblique muscles.  2) US: right rectus hemorrhage, free liquid in the right lower quadrant. CT: defect of the right anterior abdominal wall, with a slight separation between the right rectus and the oblique muscules. | 1) Surgical (urgent)  2) Surgical (urgent) | 1) SH. Omentum identified under the skin. Primary repair (absorbable sutures).  2) Exploratory laparoscopy. Peritoneal repair (partial report) | 1) Favorable  2) Favorable |
| Sinopidis et al. (2018) [75] | Greece | Newborn | Male | Left | Bilateral inguinal hernias | - | - | - | US: preperitoneal fat protruding through a defect of the transversalis fascia | Surgical | 2-month-old: bilateral inguinal hernia repair  5-month-old: SH. Preperitoneal fat adhered to the tip of the hernial sac. Primary repair. | Favorable (18m follow-up) |
| Sengar et al. (2018) [76] | India | 1) 12y  2) 4y  3) 4y  4) 2.5y  5) 2.3y  6) 2y  7) 1.6y  8) 1m  9) 1m  10) 6d | 1) Female  2) Female  3) Male  4) Male  5) Male  6) Male  7) Male  8) Female  9) Male  10) Male | 1) Bilateral  2) Right  3) Left  4) Bilateral  5) Right  6) Right  7) Right  8) Left  9) Right  10) Left | 5) Right UDT  6) Right UDT  7) Right UDT  9) Umbilical hernia, lumbar hernia  10) Hypospadias | - | - | - | - | Surgical | 3) SH containing small bowel  5,6,7,10) SH containing testis. All testes were normal without epidydimal-testicular dissociation. In all cases orchidopexy was performed. 2 cases could be brought down up to a high scrotal  level only  All cases: primary repair. | Favorable (2m-8y follow-up) |
| Fai-So et al. (2018) [77] ***** | Australia | 10y | Male | Left | - | Incarcerated sigmoid colon | Previous local trauma (bicycle handlebar) | - | CT: SH with a loop of sigmoid colon | Surgical (urgent) | Exploratory laparoscopy. Primary repair (laparoscopic). Absorbable sutures. | Favorable (5w follow-up) |
| Vega-Mata et al. (2019) [78] | Spain | 13y | Male | Right | - | Obesity (BMI 32.5) | - | - | US: 7mm SH with fat herniation | Surgical | SH. Peritoneal sac containing omentum. Primary repair (laparoscopic). Non-absorbable sutures. | Favorable (1y follow-up) |
| Deshmukh et al. (2019) [79] | India | 11m | Male | Left | Left UDT | - | - | - | - | Surgical | SH. Laparoscopic repair: Peritoneal sac present containing left testis. Orchidopexy and primary repair | - |
| Nagara et al. (2020) [80] | Japan | Newborn | Male | Left | Right-sided inguinal hernia. Left UDT. ZC4H2 associated disorders: Congenital contractures of  upper and lower extremities, hypokinesia, paraesophageal hiatal hernia | Small bowel incarceration | - | - | CT: left SH with incarcerated small bowel. Right-sided inguinal hernia. | Conservative | - | Death (SH incarceration, sepsis) |
| Taha et al. (2021) [81] | Saudi-Arabia | 50d | Male | Right | Right UDT | Incarceration | - | - | Plain abdominal X-ray: distended bowel  loops, gasless lower abdomen, and right lower quadrant lucency. US: small amount of intraperitoneal free fluid and a loop of bowel  herniated through the abdominal wall, defect; the defect was (10 mm) in diameter. | Surgical (urgent) | SH. Peritoneal sac present containing fluid, small bowel loops, and the right testis. Reduction, orchidopexy, primary repair | Complication: scrotal infection (conservative management). Testicular atrophy (2y follow-up) |
| García-Sanchez et al. (2021) [82] ***** | Spain | 11y | Female | Right | - | - | Previous local trauma (bicycle handlebar) | 2.4x1.8 cm | US: 2.4x1.8 cm defect between rectus abdominis and oblique/transversus with omentum and small bowel loops inside | Surgical | - | Favorable (1w follow-up) |
| Sinacer et al. (2021) [83] | Algeria | Newborn | Male | Right | Right inguinal hernia (incarcerated), bilateral UDT, polydactyly (right hand), anal stenosis (type 1 diabetic mother) | - | - | 1.5 cm | - | Surgical | SH. Peritoneal sac present containing small bowel loops and the left testis. Reduction, orchidopexy, primary repair (non-absorbable sutures) | - |
| Tahmri et al. (2021) [84] | Tunisia | 9y | Male | Right | - | - | Previous local trauma (bicycle handlebar) | 2 cm | US: right rectus abdominis hematoma. Diastasis between the lateral edge of the rectus abdominis and the ipsilateral oblique and transverse muscles resulting in a 14 mm hernia sac containing omentum | Surgical | SH. Peritoneal sac present. Primary repair (absorbable sutures) | Favorable |
| Gonuguntlaet al. (2022) [85] | India | 3m | Male | Left | Left UDT | - | - | - | US: 2 abdominal wall  defects. 1) in the left iliac fossa (1.6 cm)  2)  posterosuperior to the  the first defect in the posterior abdominal wall lateral to the kidney  (1.3 × 0.6 cm). Left testis not visible. | Surgical | SH. Laparoscopic repair. Absence of inguinal canal and gubernaculum.Orchidopexy and primary repair | Favorable (1y follow-up) |
| Kropilak et al. (2022) [86] | USA | 8y | Male | Left | - | - | Previous local trauma (bicycle handlebar) | - | CT: traumatic SH in the left lower quadrant with incarceration of loops of small bowel and associated stranding of surrounding tissues. | Surgical (urgent) | Diagnostic laparoscopy. Laparotomy conversion. SH. Small bowel mesenteric injury with a devitalized jejunum. Intestinal resection and anastomosis. Primary repair (absorbable suture) |  |
| Okumus et al. (2022) [87] | Turkey | Newborn | Male | Right | Right UDT | - | - | 2-3 cm | Plain abdominal X-ray: intestinal loops under the skin, US: Ventral hernia | Surgical | SH. Peritoneal sac present containing the right testis. Orchidopexy, primary repair. | Favorable (6m follow-up) |
| Kangabam et al. (2023) [88] | India | 17y | Male | Right | - | - | Previous local trauma (motorcycle handlebar) | 1 cm | US: 1x1 cm defect in right Spigelian aponeurosis with herniating bowel loops  CT: confirmation of findings | Conservative (patient´s choice) | - | - |
| Farina et al. (2024) [89] | Italy | 4m | Male | Left****** | Left UDT | Large Bowel Incarceration | - |  | US: 5,8 mm hernia breach containing a sac with colon loop, adipose tissue, fluid collection and a testicle. | Surgical (urgent) | Herniopexy and testicle repositioning in the scrotal sac | Favorable |
| Ablatt et al. (2025) [90] | USA | 2w | Male | Left | Left UDT, bilateral inguinal hernia, umbilical hernia | - | - |  | US: left inguinal hernia containing fluid and nonobstructive bowel. SH not diagnosed. | Surgical (urgent) | Open umbilical hernia repair, laparoscopic left orchiopexy, open SH repair, open left inguinal hernia repair, open right inguinal hernia repair | Favorable (6w follow-up) |

**Table 1. Previously reported cases of pediatric spigelian hernia in the medical literature**

SH: Spigelian Hernia; US: ultrasound; NR: not reported; y: years; m: months; d: days; w: weeks; USA: United States of America; UK: United Kingdom; cm: centimeters; ESR: elective surgical repair; In some cases, there was duplication or redundancy of patients between publications: CT: computer tomography; US: ultrasound; IC: intercostal; UDT: undescended testis; TAWH: Traumatic abdominal wall hernia; BMI: Body mass index.

*The original article could not be found, and the data reported correspond to other previous reviews; **: the histological study showed muscle atrophy with fatty infiltration; ***: authors suspect that it was an incisional hernia (postoperative hernia); ****: Siblings; *****: reported unspecifically as a handlebar hernia or traumatic abdominal wall hernia (TAWH) but radiologically and clinically compatible with an SH, ******: The authors report inconsistent data in the manuscript, sometimes referring to left-sided laterality and other times to right-sided laterality. However, the overall interpretation of the case suggests that it involves left-sided laterality. The authors were contacted to resolve this matter.
