## Supplementary File 1. for "Pediatric Spigelian Hernia and Spigelian-Cryptorchidism Syndrome: an Integrating Review"

**Supplementary File 1. Inclusion and Exclusion Criteria**

**Inclusion criteria**

-Prospective or retrospective original studies, case series, or individual cases reporting patients with a diagnosis of Spigelian hernia.

**Exclusion criteria**

-Duplicate or overlapping studies.

-Reviews, systematic reviews, consensus guidelines.

-Languages other than English or Spanish.

-Studies with no population of interest.

-Studies conducted in adult patients.
